# Prediction of the Peak, Effect of Intervention and Total Infected by the Coronavirus Disease in India

**DOI:** 10.1101/2020.04.20.20072793

**Authors:** Parth Vipul Shah

## Abstract

We study the effect of the coronavirus disease 2019 (COVID-19) in India using the SEIR compartmental model. After it’s outbreak in Wuhan, China, it has been imported to India which is a densely populated country. India is fighting against this disease by ensuring nationwide social distancing. We estimate the infection rate to be 0.258 using a least square method with Poisson noise and estimate the reproduction number to be 2.58. We approximate the peak of the epidemic to be August 11, 2020. We estimate that a 25% drop in infection rate will delay the peak by 38 days for a 1 month intervention period. We estimate that the total individuals infected in India will be approximately 9% of the total population.

## I. INTRODUCTION

In this work, on the eve of the outbreak of the deadly coronavirus disease 2019 (COVID-19)^1^ in India, caused by severe acute respiratory syndrome coronavirus 2 (SARS-CoV-2)^2^, we predict the peak of the epidemic, effect of intervention and total individuals infected. This disease has demonstrated person-to-person transmission^3^ and as India is a densely populated country, it is necessary to understand the effects of this disease to mitigate its risk. January 30, 2020 marked the first identified case of COVID-19 in India.^4^ Meanwhile COVID-19 has spread around the world. As of March 22, 2020, Italy has approximately 59 thousand confirmed cases, USA has approximately 34 thousand confirmed cases and Spain has approximately 29 thousand confirmed cases. India has only a total of 66 confirmed cases.^4^ The World Health Organization (WHO) declared the disease a pandemic on March 11, 2020.^5^ The total number of confirmed cases in India and the daily increase in cases is depicted below in Figures 1 and 2.

**FIG. 1:**
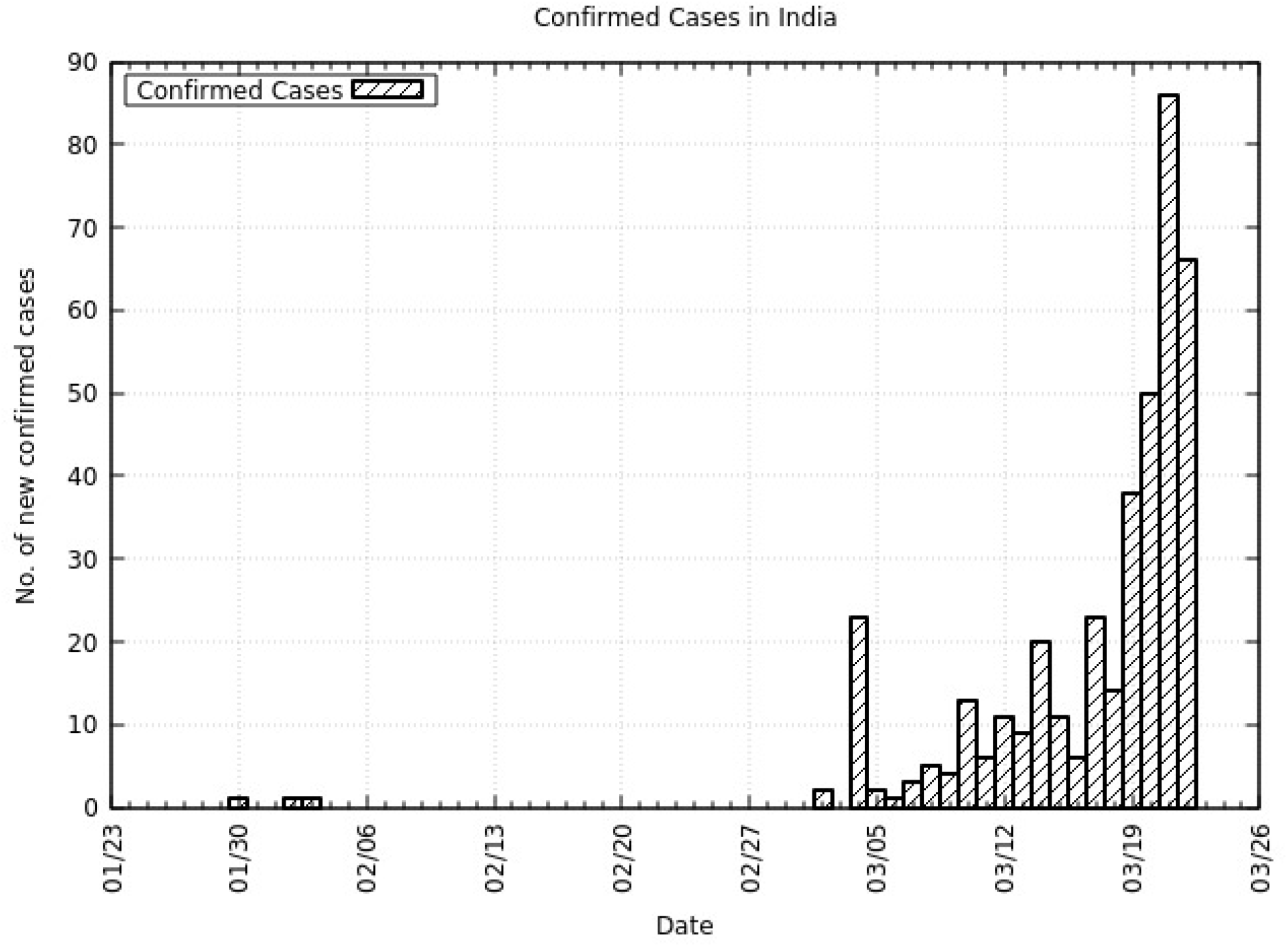
Daily increase in confirmed cases of COVID-19 in India. Day 0 is January 22, 2020 and day 52 is March 22, 2020. Data is taken from Center for Systems Science and Engineering (CSSE) at Johns Hopkins University (JHU).^4^

**FIG. 2:**
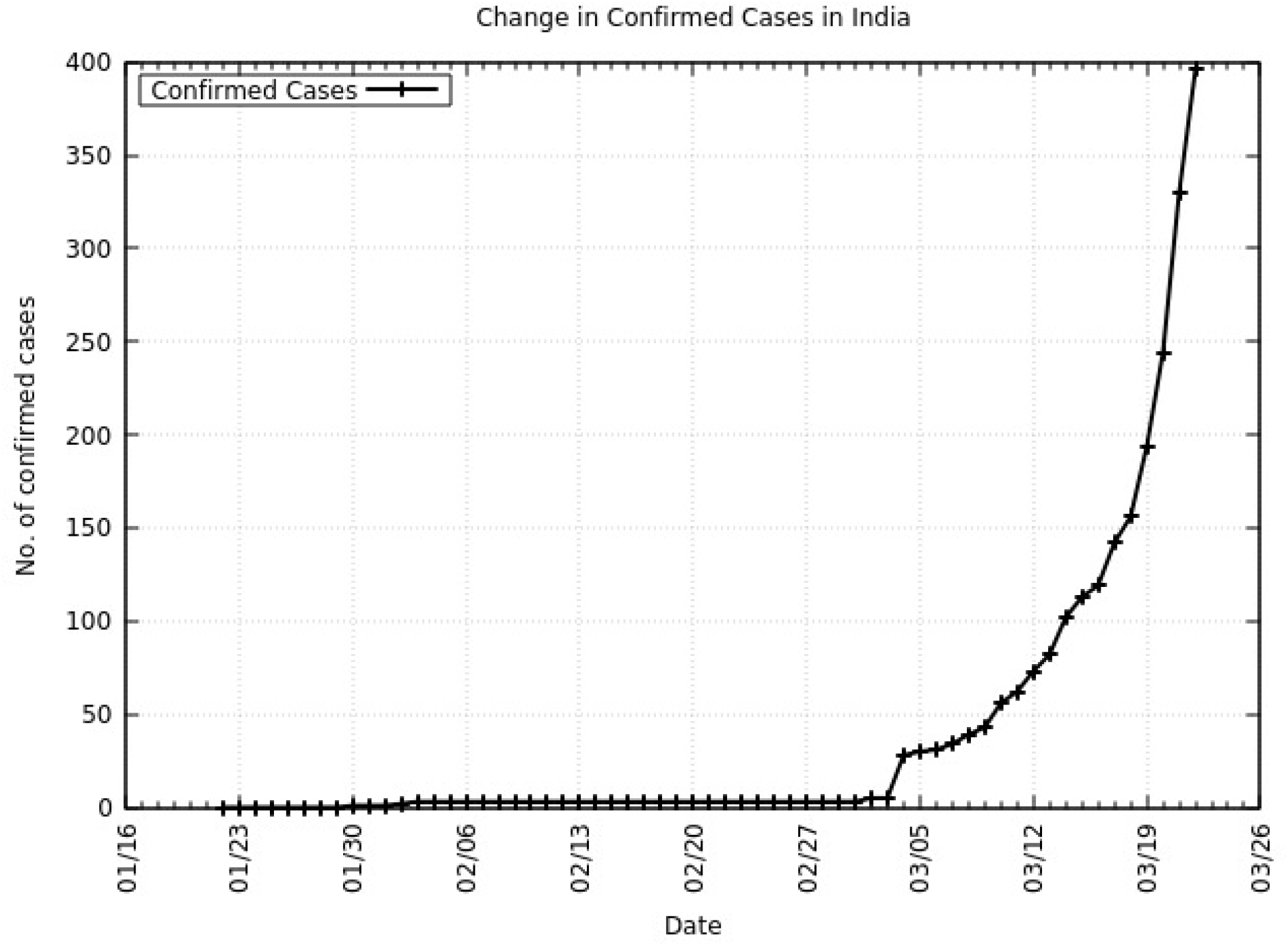
Cumulative confirmed cases of COVID-19 in India. Day 0 is January 22, 2020 and day 52 is March 22, 2020. Data is taken from Center for Systems Science and Engineering (CSSE) at Johns Hopkins University (JHU).^4^

The growth of the number of confirmed COVID-19 cases can be approximated by an exponential function. The growth factor *r* at day *t* can be calculated as

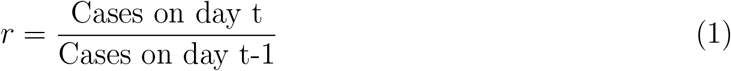

March 22, 2020, India is reporting an average growth rate of 1.916 calculated over the past 4 days (refer to Table I). Stricter rules and measures of ensuring social distancing have begun.^6^ India is fighting this disease by ensuring nation-wide lock-down and social distancing when the number of infected individuals are low. All non-essential workers across all industries have been given stay at home orders.

**TABLE I:** Growth Factor, r

| Date | Day | No. of confirmed cases | Growth factor, $r$ |
| --- | --- | --- | --- |
| March 18 | 48 | 14 | - |
| March 19 | 49 | 38 | 2.714 |
| March 20 | 50 | 50 | 1.315 |
| March 21 | 51 | 86 | 1.720 |

## II. METHODS

### A. Model

The SEIR compartmental model has been used to model previous pandemics in the field of Epidemiology. It is applied on the data from India based on a recent publication^7^ which applied it on the data from Japan. The same methodology is adopted here. The assumption that once an individual contracts the virus and recovers, the individual is immune to this virus is assumed. The SEIR model is defined in Equation 2.

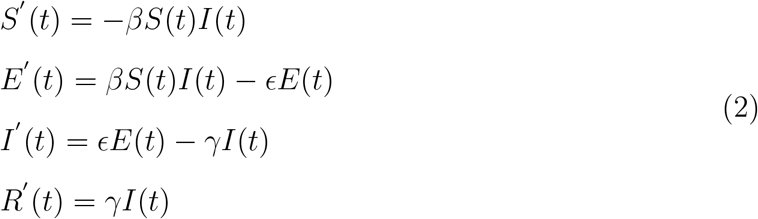

The susceptible, exposed, infected and removed populations at time t is denoted by *S*(*t*), *E*(*t*), *I*(*t*), *R*(*t*) respectively. The infection rate, the onset rate and the removal rate is denoted by *β, ϵ, γ* respectively. The average incubation period and the average infectious period is 1*/ϵ* and 1*/γ* respectively. We fix 1*/ϵ* to 5 and 1*/γ* to 10 based on recent publications that have studied the average incubation period and the average infectious period.^89^ The unit time is 1 day. The fraction of infected individuals that can be identified by diagnosis is denoted by *p*. We also fix *S* + *E* + *I* + *R* to 1 so that calculations are a proportion of the total population. The total population of India, N is fixed at 1.33 billion.^10^ The assumption that 1 individual amongst a total population of N (1339.2 × 10^6^) individuals is identified as infected at *t* = 0 is adopted. Therefore the total number of infected individuals who are identified at time t is given by the product of fraction of individuals that can be identified by diagnosis, infected individuals at time t and the total population (Equation 3).

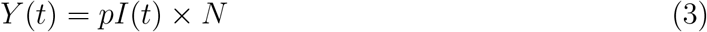

To obtain the initial conditions of the model, we assume that there are no exposed or removed populations at *t* = 0.

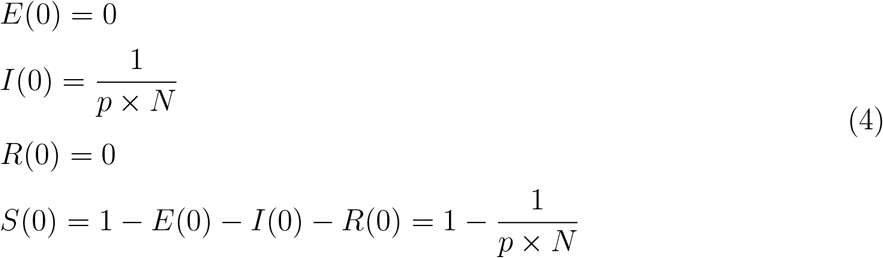

are the initial conditions. We fix 0.01 *< p <* 0.1 based on government reports of the density of affected people in a particular region.^11^ The expected value of secondary cases produced by one infected individual, the reproduction number, *R*_0_ is calculated from Equation 5.^12^

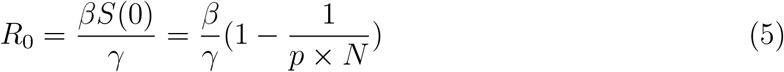

Using this model, we estimate the infection rate, *β* in the next section.

### B. Estimation of the infection rate, *β*

Let *y*(*t*), *t* = 0, 1, 2…52 be the daily confirmed cases of COVID-19 in India from January 22, 2020 (*t* = 0) to March 22, 2020 (*t* = 52). Using the least square approach with Poisson noise to estimate the infection rate, the following steps are adopted. With Poisson noise, Equation 3 is modified to

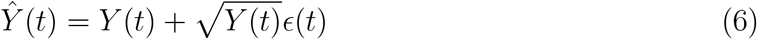

*ϵ*(*t*), *t* = 0, 1, 2…52 are random variables from a standard normal distribution. The following steps are adopted to estimate *β*.

1. For *β >* 0, calculate *Y* (*t*), *t* = 0, 1, 2…52 using Equation 3.
2. Calculate *Ŷ* using Equation 6.
3. Calculate 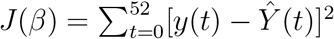
4. Run step 1 to step 3 for 0.2 ≤ *β* ≤ 0.4 and find *β*\* such that *J* (*β*\*) = min_0.2≤*β*≤0.4_*J* (*β*)
5. Repeat step 1 to step 4, 10000 times and obtain the distribution of *β*\*. Approximate the same by a normal distribution and obtain the 95% confidence interval.

We obtain a value of *β* equal to 0.258 and the 95% confidence interval as 0.250 *−* 0.266. Also, *R*_0_ is equal to 2.58 and the 95% confidence interval as 2.50*−*2.66. This is represented in Figure 3 with an overlap of the number of new confirmed cases. A summary of all parameters can be found in Table II.

**TABLE II:** Parameters

| Parameter | Description | Value |
| --- | --- | --- |
| $\beta$ | Infection rate | 0.258 |
| $R_0$ | Reproduction number | 2.58 |
| $\epsilon$ | Onset rate | 0.2 |
| $\gamma$ | Removal rate | 0.1 |
| $N$ | Total population of India | $1339.2 \times 10^6$ |
| $p$ | Identification rate | $0.01 - 0.1$ |

**FIG. 3:**
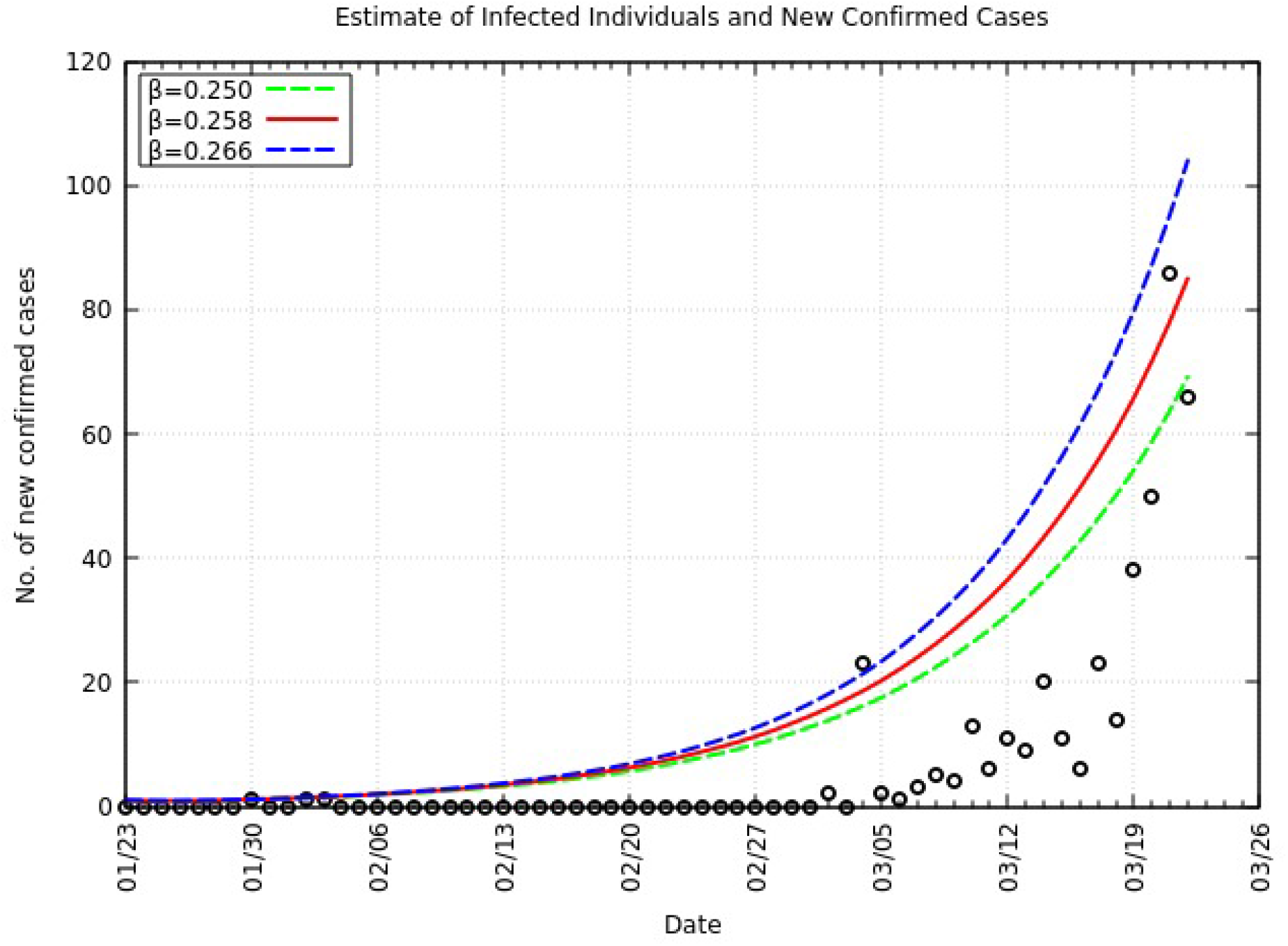
Comparison of daily confirmed cases and *Y* in India from *t* = 0 to *t* = 52

## III. RESULTS

### A. Peak Prediction

The epidemic peak, *t*\* is defined as the day t when the maximum value *Y* in a period of 550 days is achieved. As an expression, *Y* (*t*\*) = max_0≤*t*≤550_. As the epidemic peak and size are sensitive to the identification rate p, we report the following.

For *p* = 0.1, the estimated peak is *t*\* = 229 with a 95% confidence interval of 221-238. That is, starting from January 22, 2020 (*t* = 0), the estimated peak is September 7, 2020 (*t* = 229) and interval ranging from August 30, 2020 to September 16, 2020. This is represented in Figure 4.

**FIG. 4:**
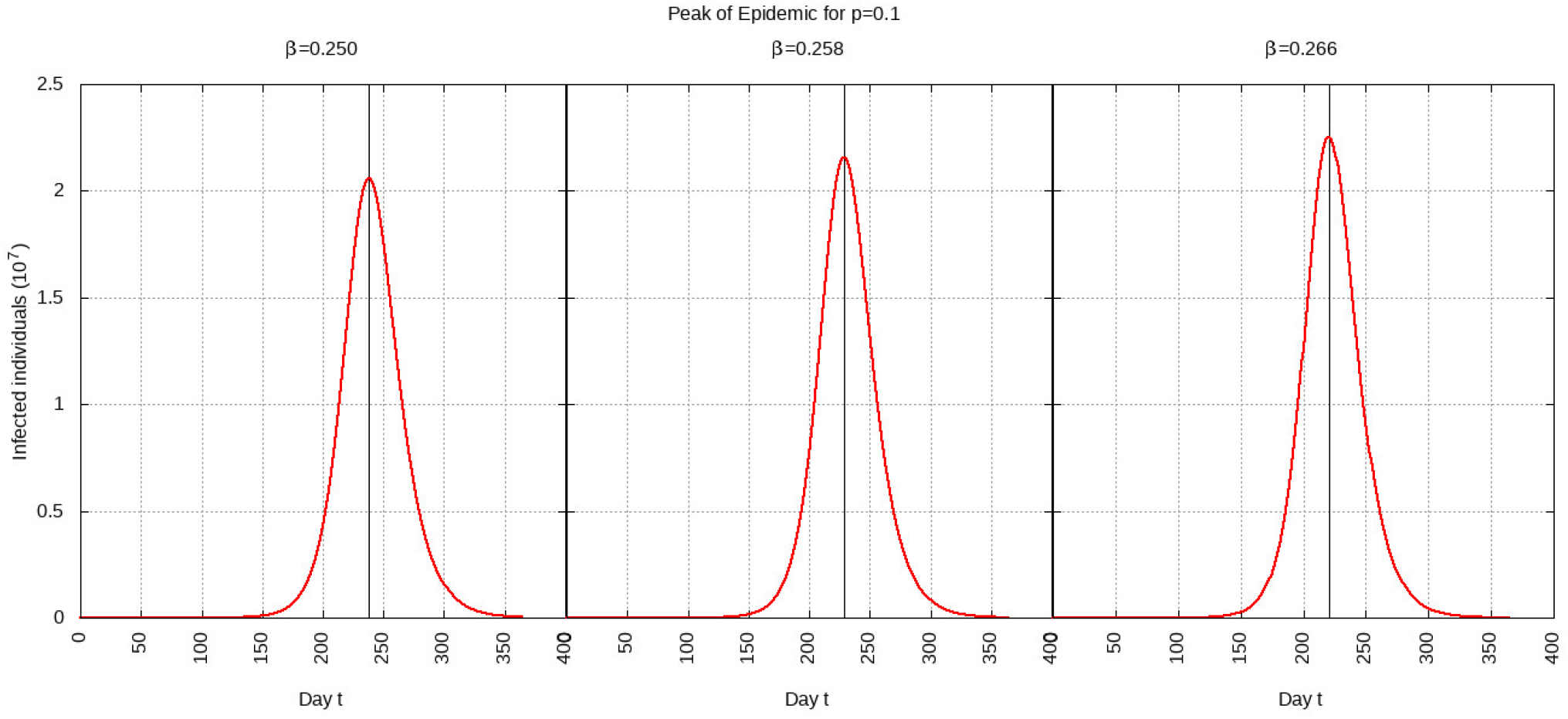
Infected individuals for time t, 0 ≤ *t* ≤ 550 for *p* = 0.1

For *p* = 0.01, the estimated peak is *t*\* = 202 with a 95% confidence interval of 194-210. That is, starting with January 22, 2020 (*t* = 0), the estimated peak is August 11, 2020 (*t* = 202) and interval ranging from August 3, 2020 to August 19, 2020. This is represented in Figure 5.

**FIG. 5:**
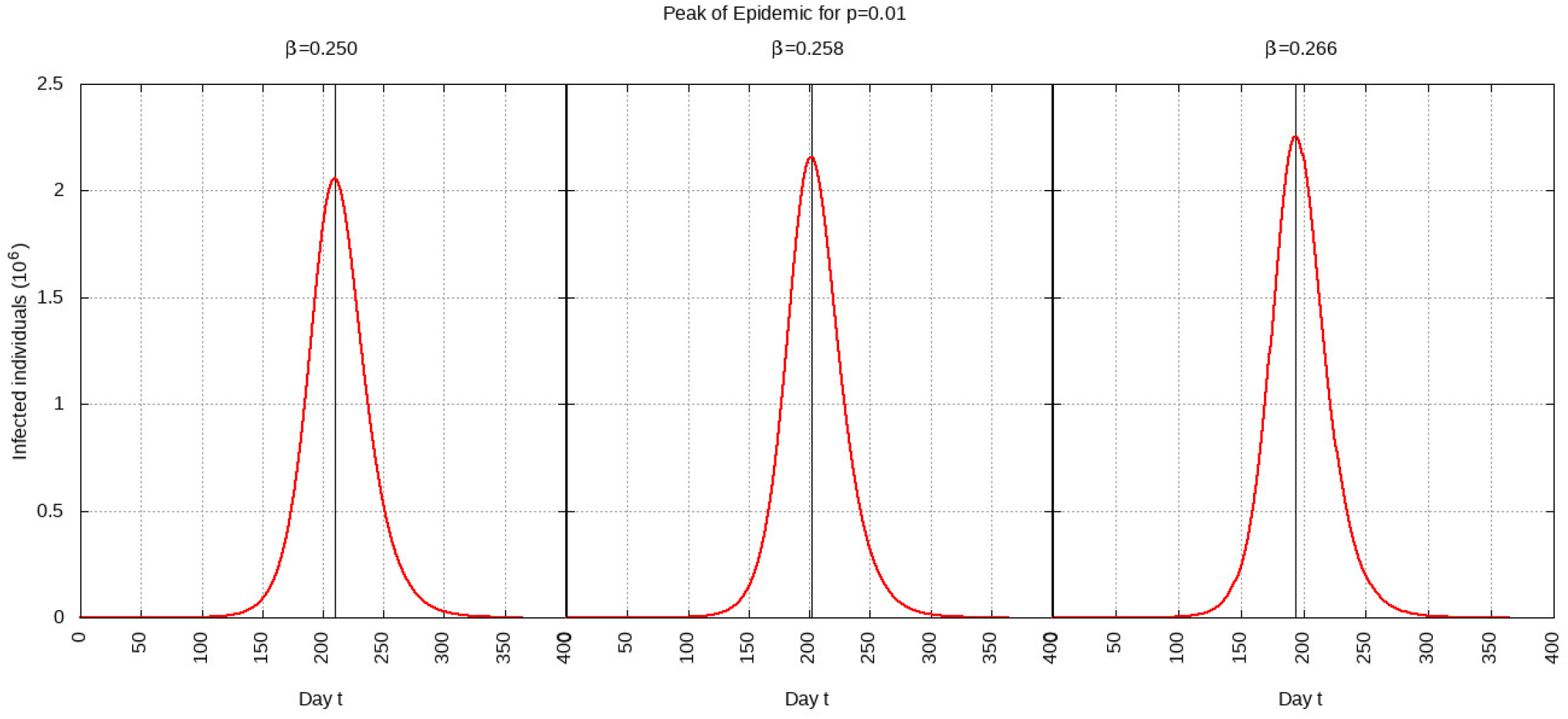
Infected individuals for time t, 0 ≤ *t* ≤ 550 for *p* = 0.01

### B. Effect of Intervention

India has announced a 21 day complete shutdown starting March 25, 2020.^13^ For our calculations, we assume that this shutdown will reduce the infection rate to 75%, 50% or 25% of its original value of 0.258. For each reduction in infection rate, the effect of an intervention period of 30 days (one month) and 180 days (six months) is calculated. The government of India maintains that the third stage or community transmission has not begun.^14^ The current identification rate is low due to the low number of COVID-19 tests being conducted.^15^ Therefor, we set p to 0.01 as the current identification rate is low. This closely matches available data too.

#### 1. 75% Reduction in Infection Rate

For a 75% reduction in the infection rate and one month of intervention, the estimated peak is delayed by 12 days. That is, pushed from 202 days to 214 days since day 0 (January 30, 2020). For a 75% reduction in the infection rate and six months of intervention, the estimated peak is delayed by 68 days. That is, pushed from 202 days to 270 days since day 0 (January 30, 2020). These are represented in Figure 6.

**FIG. 6:**
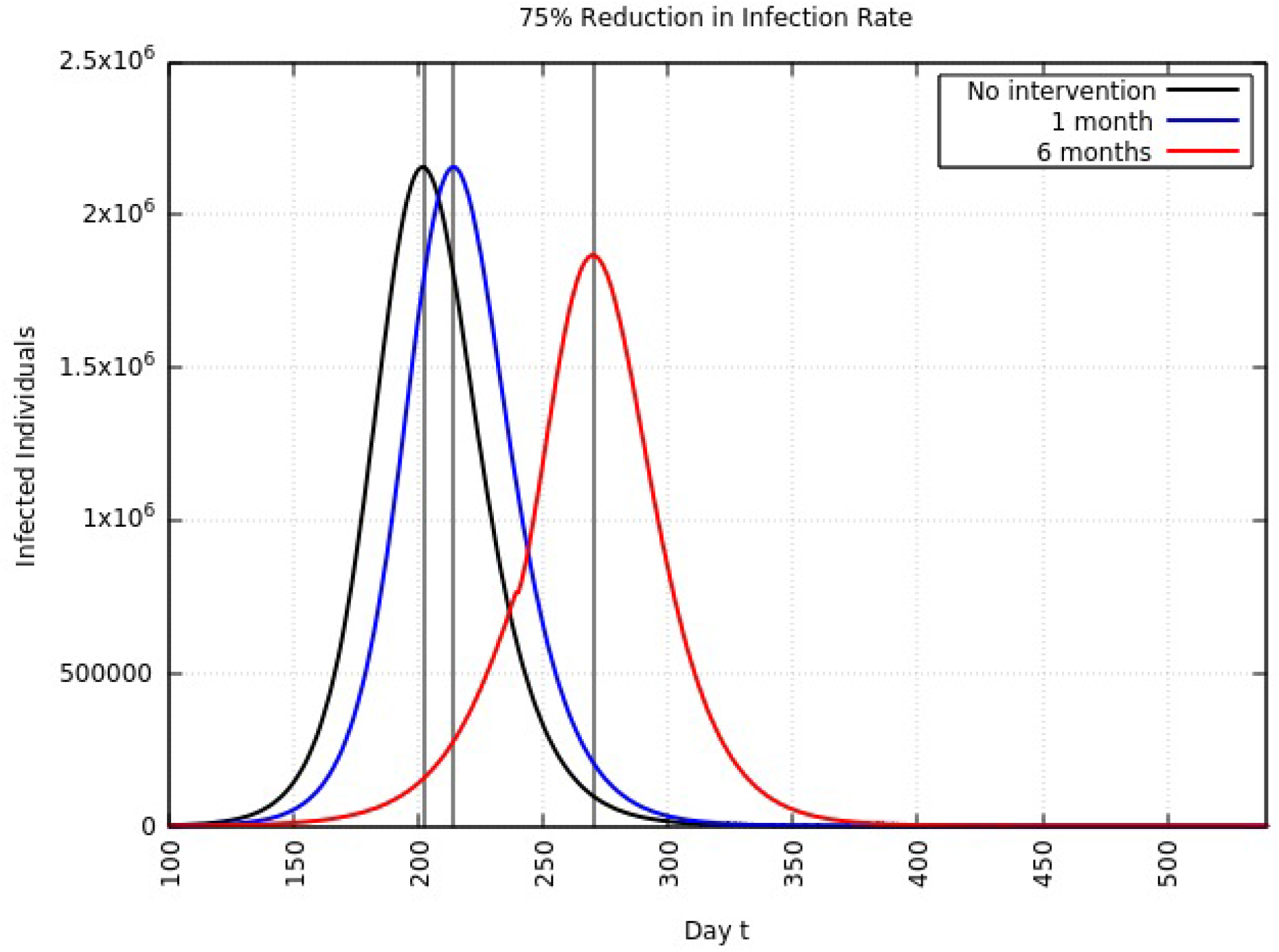
Variation in *Y* (*t*) for time t, 0 ≤ *t* ≤ 550 with no intervention, 1 month of intervention and 6 months of intervention with assumption of *β* = 0.75 × 0.258

#### 2. 50% Reduction in Infection Rate

For a 50% reduction in the infection rate and one month of intervention, the estimated peak is delayed by 24 days. That is, pushed from 202 days to 226 days since day 0 (January 30, 2020). For a 50% reduction in the infection rate and six months of intervention, the estimated peak is delayed by 141 days. That is, pushed from 202 days to 343 days since day 0 (January 30, 2020). This is represented in Figure 7.

**FIG. 7:**
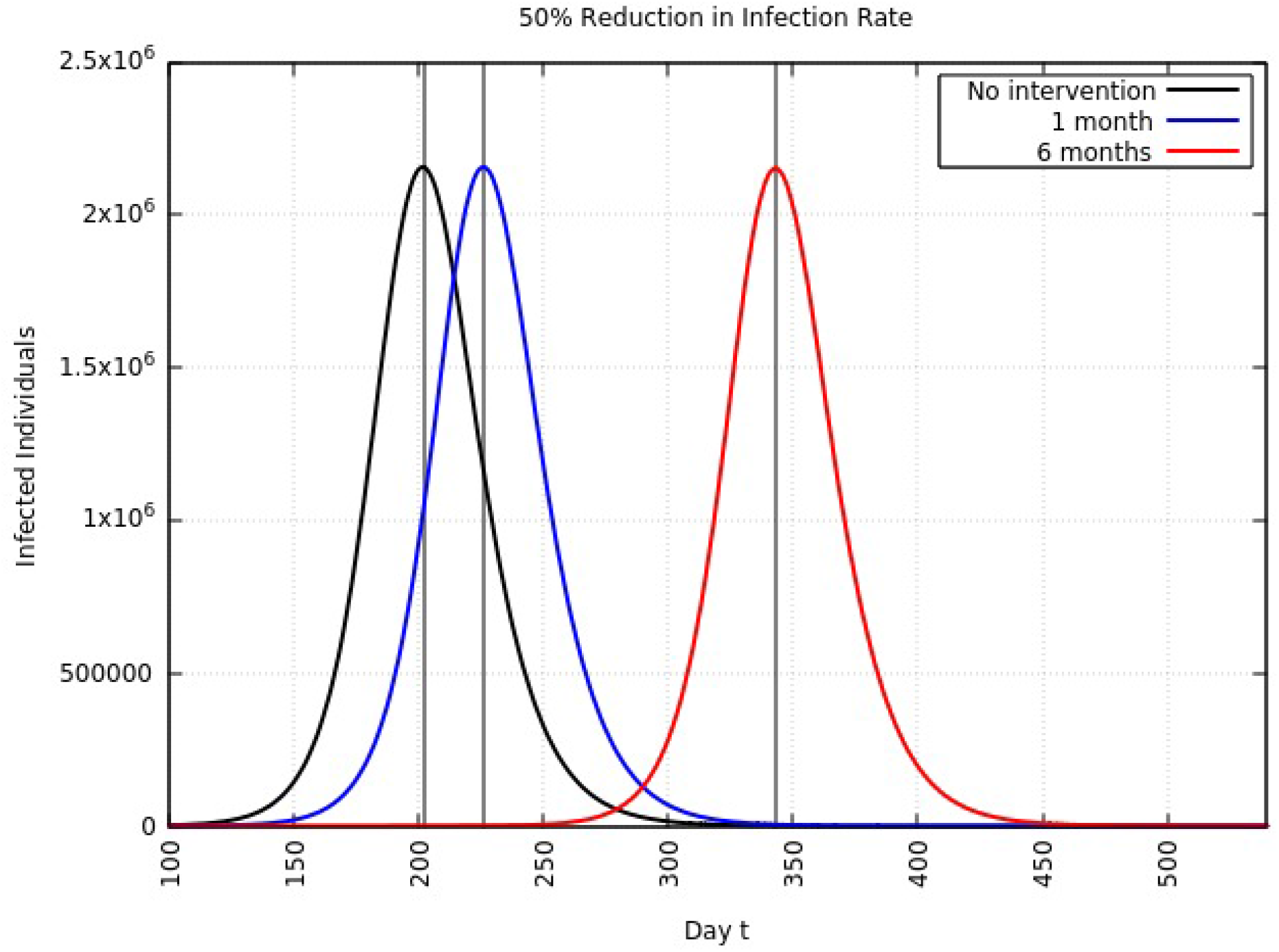
Variation in *Y* (*t*) for time t, 0 ≤ *t* ≤ 550 with no intervention, 1 month of intervention and 6 months of intervention with assumption of *β* = 0.50 × 0.258

#### 3. 25% Reduction in Infection Rate

For a 25% reduction in the infection rate and one month of intervention, the estimated peak is delayed by 38 days. That is, pushed from 202 days to 240 days since day 0 (January 30, 2020). For a 25% reduction in the infection rate and six months of intervention, the estimated peak is delayed by 234 days. That is, pushed from 202 days to 436 days since day 0 (January 30, 2020). This is represented in Figure 8.

**FIG. 8:**
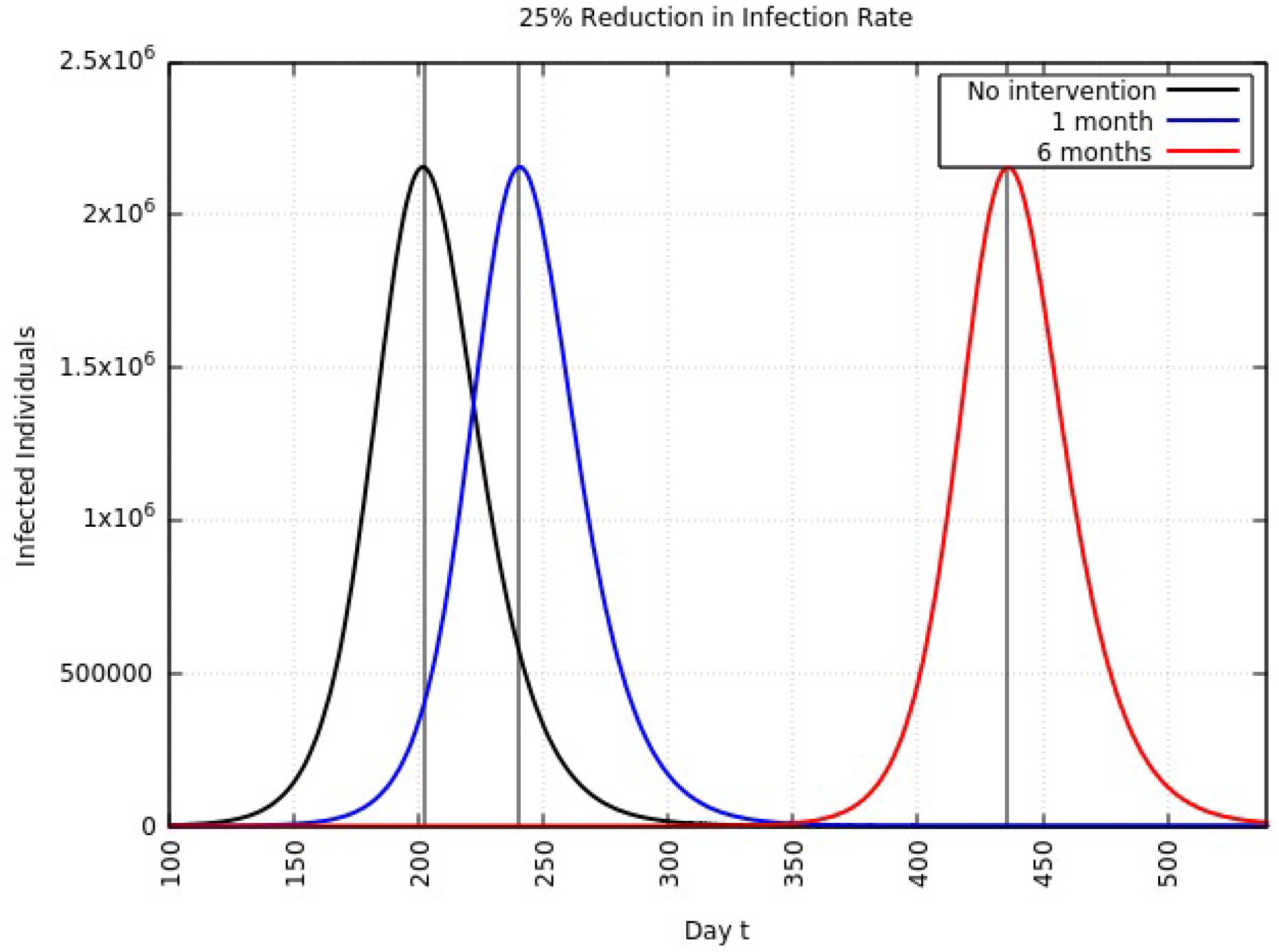
Variation in *Y* (*t*) for time t, 0 ≤ *t* ≤ 550 with no intervention, 1 month of intervention and 6 months of intervention with assumption of *β* = 0.25 × 0.258

A summary of all the delays in the estimated peaks can be found in Table III.

**TABLE III:** Summary of change in estimated peak from August 11, 2020. Units are days. The 1 month and 6 months delays are the delays in the estimated peaks after respective intervention periods. The dates indicated are the revised dates of the estimated peaks.

| $\beta$ Drop | 1 Month Delay | Date | 6 Months Delay | Date |
| --- | --- | --- | --- | --- |
| 75% | 12 | August 23 | 68 | October 18 |
| 50% | 24 | September 4 | 141 | December 30 |
| 25% | 38 | September 18 | 234 | April 2, 2021 |

### C. Total Individuals Infected

Upon integrating *Y* (*t*) from Equation 3 over time *t*, we obtain an approximate of the total number of infected individuals over 550 days. With no intervention, the approximate total number of infected individuals will be 1.2087 × 10^8^ which is 9% of the total population of India. With one month of intervention and 75% reduction in the infection rate, the approximate total number of infected individuals will be the same. But with six months of intervention and 75% reduction in the infection rate, the approximate total number of infected individuals will be 1.2002 × 10^8^ compared to 1.2087 × 10^8^ with no intervention. This means approximately 850,000 individuals will not be infected. The change in these numbers for a higher reduction in infection rate does not significantly change the approximate total number of infected individuals. It only pushes the approximate epidemic peak to a later date.

## IV. DISCUSSION

From this work, by applying the SEIR compartmentalization model, it is clear that the COVID-19 epidemic peak can easily reach August, 2020. This prediction is sensitive to changes in the behavior of not only the virus but of people and their practice of social distancing. India is still in the process of gearing up its healthcare system and it currently may be ill equipped to deal with a large number of cases. Delaying the peak will ensure adequate medical equipment and personnel for all her citizens. The WHO has issued similar statements reiterating the importance of practicing social distancing to ensure that national health care systems are not strained.

## Data Availability

All data referred to in the manuscript is publicly available.

## Acknowledgements

The author would like to thank anonymous reviewers for their inputs

## Conflicts of Interests

The author declares no conflict of interest

## Declarations of Interest

None

## Funding

None

## Notes

### Competing Interest Statement

The authors have declared no competing interest.

### Funding Statement

No external funding was received.

